# Therapeutic delivery from a solid formulation during breastfeeding: a feasibility study

**DOI:** 10.1101/2020.03.05.20031583

**Authors:** Theresa Maier, Paula Peirce, Laura Baird, Sophie L. Whitehouse, Nigel K. H. Slater, Kathryn Beardsall

**Affiliations:** Department of Chemical Engineering and Biotechnology, University of Cambridge, West Cambridge Site, Philippa Fawcett Drive, Cambridge CB3 0AS, UK; Department of Paediatrics, University of Cambridge, Biomedical Campus, Cambridge CB2 0QQ, UK; Cambridge University Hospitals NHS Foundation Trust, Biomedical Campus, Cambridge CB2 0QQ, UK; College of Social Sciences, Arts and Humanities, University of Leicester, Leicester, LE2 1RQ, UK

**Keywords:** Breastfeeding, human milk, oral therapeutic delivery, medication systems, newborn infant, preterm infant, neonatal intensive care

## Abstract

**Background:** Enteral drug and nutrient delivery to breastfed infants depends on the use of oral syringes and liquid formulations. This can pose both practical and emotional challenges to drug delivery.

**Objectives:** The presented study aimed to explore the potential of using solid formulations for therapeutic delivery during breastfeeding.

**Methods:** Single centre feasibility study within a tertiary level neonatal unit in the UK, involving twenty-six breastfeeding mother-infant dyads. Vitamin B12 was delivered to infants during breastfeeding from a solid formulation within a commercial silicon nipple shield. Outcomes included the quantitative measurement of change in serum vitamin B12 and a mixed methods assessment of maternal expectations and experiences.

**Results:** Participants described the surprising ease of ‘drug’ delivery, with no negative impact on breastfeeding behaviour or sensation reported. Vitamin B12 levels rose on average from a baseline of 533 pg/mL (236 - 925 pg/mL) to 1871 pg/mL (610 – 4981 pg/mL) at 6 - 8 hours post-delivery. All mothers expressed their support for this approach, 85% a preference over the use of oral syringes. Reasoning for support related to the reduced medicalisation of this procedure compared to the use of oral syringes, and a desire for choices in relation to their infants’ health.

**Conclusions:** This study demonstrated that therapeutic delivery from a solid formulation within a nipple shield was feasible and acceptable to mothers and infants.

## INTRODUCTION

Drug and nutrient delivery to breastfed infants can be challenging for parents, as many babies demonstrate aversive behaviour towards drug delivery from oral syringes.[1] In low-resource settings, additional challenges of liquid formulations arise due to the lack of refrigerated storage.[2] To address this shortcoming, research at the University of Cambridge’s Department of Paediatrics and Department of Chemical Engineering and Biotechnology has been exploring alternative approaches to therapeutic delivery in infancy. *In-vitro* studies have previously demonstrated the potential of drug administration using a breastfeeding simulation apparatus. [3,4] Qualitative studies within our Neonatal Unit also demonstrated parents’ and nursing staff’s support for such an intervention, indicating that it could help foster mother-infant bonding and encourage parental empowerment.[2]

The study aimed to evaluate the feasibility and acceptability of solid formulation delivery to infants while breastfeeding. Following previous lab-based and qualitative research, it is the first clinical feasibility study to ever be undertaken with mother-infant dyads.

## METHODS

### Study design

This was a single centre feasibility study, conducted from July to November 2018 at the University of Cambridge Addenbrooke’s Hospital Trust. The intervention involved the delivery of vitamin B12 to breastfed infants via a nipple shield during a single breastfeed. Assessments included quantitative measurements of serum vitamin B12 levels at baseline and 6 - 8 hours after the intervention, as well as a mixed methods assessment of maternal expectations, experience, and acceptability. The latter was conducted by the first author, a single female researcher with clinical research training. A qualified nurse or lactation consultant known to the mother provided breastfeeding support and advised on the appropriate application of the nipple shield to the breast.

### Study population and participant recruitment

Breastfeeding mother-infant dyads were recruited through convenience sampling from the neonatal unit and postnatal transitional care wards.

The study’s intention and practice was to recruit infants on the postnatal wards below one moth of corrected age, while the upper limit for inclusion included corrected age <12 months. Infants had to be clinically stable and competent at breastfeeding, as confirmed by the lactation team. Exclusion criteria were hypersensitivity to the vitamin B12 tablet’s ingredients, short bowel syndrome, and malabsorption. No restrictions related to prior B12 supplementation were applied, or potential interactions with human milk considered, as the study was solely designed as a proof-of-principle for nutrient / therapeutic delivery during breastfeeding, not to determine B12 absorption kinetics. Potentially eligible mother-infant dyads were identified by the clinical team, following which mothers were approached by the research team. The total number of participants was determined based on guidelines for feasibility studies and qualitative research, and in alignment with the objective of reaching information saturation.[5,6]

### Nipple shield and vitamin formulation

Commercial ultrathin contact nipple shields (Medela, UK) of 16 mm, 20 mm, or 24 mm size, as recommended by the lactation support team, were selected and worn by mothers during the study feed. Immediately prior to the breastfeed, a sublingual vitamin B12 tablet (1000 μg Methylcobalamin, JustVitamins Ltd, UK), suitable for vegans and without known allergenic components, was placed into the shield’s teat, following which the mother breastfed as usual. A baseline blood sample was taken prior to and a peak level at 6 - 8 hours after the study feed. The timing for collection was based on the pharmacokinetic profile and anticipated peak B12 levels.[7] Samples were immediately centrifuged and separated, with serum stored at −20°C for later batch analysis. Serum vitamin B12 levels were analysed using a LOCI vitamin B12 assay (Siemens Healthcare) at the Core Biochemical Assay Laboratory (CBAL), Cambridge University Hospitals. Vitamin B12 was chosen as a therapeutic, due to its physiological importance for infant development, and its known safety profile over a wide dosage range.[8] Within this study, it served as a model compound for other therapeutic formulations.

### Mixed methods approach

A combination of tablet-based Likert-scale questionnaires and recorded semi-structured interviews, developed based on established guidelines,[9] were undertaken before and after the study feed. Interviews were performed in a quiet area at the infants’ cot side to avoid separation of mother and infant. Discussions focused on the evaluation of maternal expectation, experience, and acceptability of the intervention. Interviews were voice-recorded, but to support honest critical reporting, mothers also provided scored answers on a tablet. Data on those scores was quantitatively evaluated, while semi-structured interviews were transcribed verbatim, and potentially identifiable data anonymized. Analysis was facilitated by ATLAS.ti (Scientific Software Development GmbH) using an inductive approach of thematic content analysis.[10,11] Hereby, an initial coding framework emerged following pre-reading, a line-by-line open-coding step, and regrouping. A final coding framework was developed through iterative revisions within the research team. Supporting quotes are used in the following tables and the main body of text to illustrate key findings, content in square brackets has been edited to improve clarity and brevity. The abbreviation “NS” and “no NS” refer to the current nipple shield use of the participating mother-infant pair, “Mx” to a mother’s participant code. Further quotes are included in the article’s supplementary data.

## RESULTS

A total of 60 infants were screened, including 4 twin pairs, of which 43 were eligible for participation. Reasons for ineligibility included: health problems or feeding difficulties (9), change to bottle feeding (4), discharge (4). Twenty-six mother-infant dyads provided their consent, of which twenty dyads completed the full study protocol (Table 1). Reasons for declining participation included: mother felt overwhelmed with the establishment of breastfeeding (3), mother did not wish to have further infant blood samples taken (3), mother declined vitamin administration (1), mother had stopped using a nipple shield (2), no reason provided (8). Reasons for non-completion/exclusion from analysis in the six cases were: change to bottle feeding (1), discharge before study feed (2), blood sampling time not kept (2), and parent-led withdrawal (1).

**Table 1:**
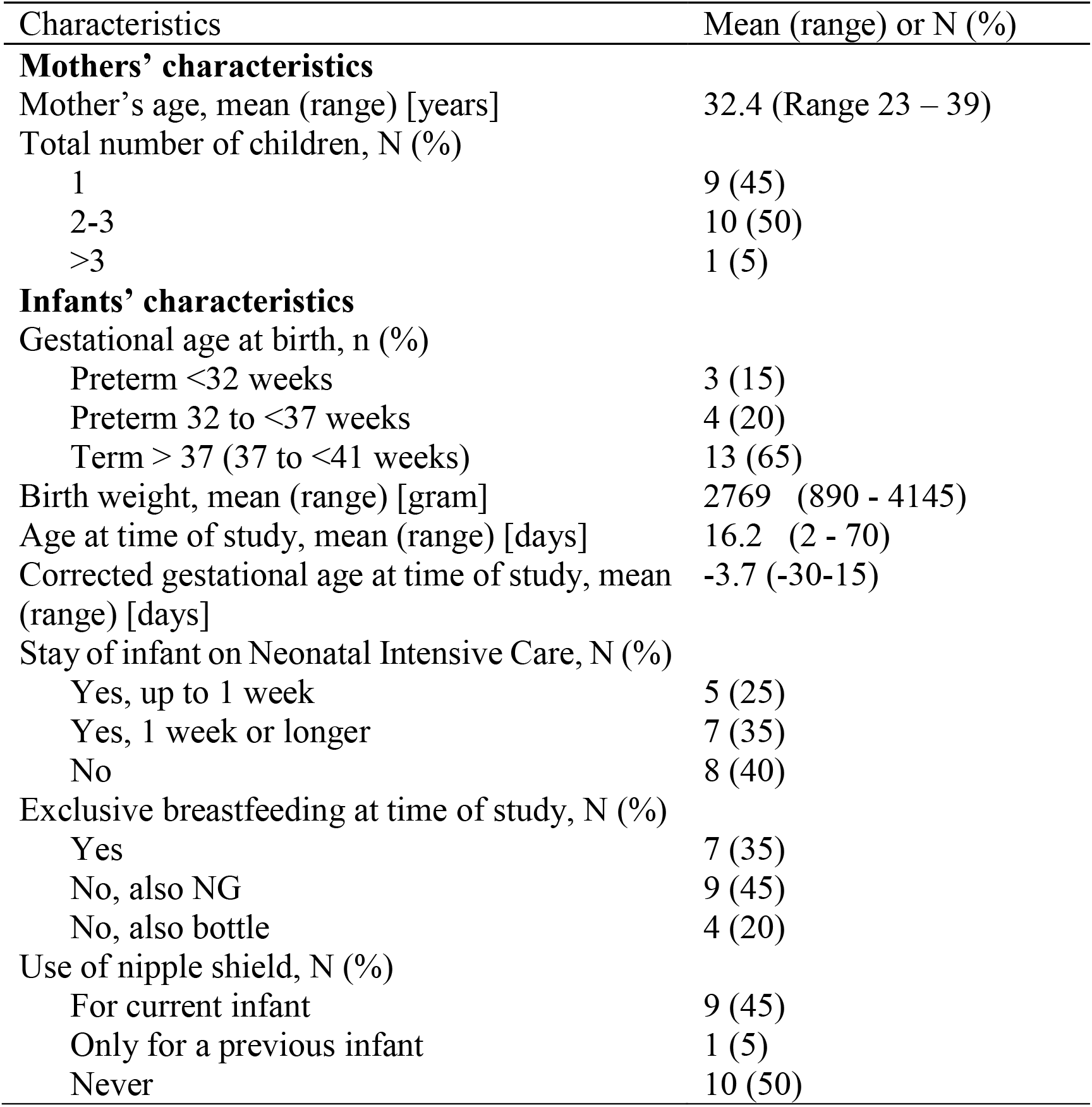
Participant characteristics (mother-infant pairs, n = 20).

| Characteristics | Mean (range) or N (%) |
| --- | --- |
| **Mothers' characteristics** |  |
| Mother's age, mean (range) [years] | 32.4 (Range 23 – 39) |
| Total number of children, N (%) |  |
| 1 | 9 (45) |
| 2-3 | 10 (50) |
| >3 | 1 (5) |
| **Infants' characteristics** |  |
| Gestational age at birth, n (%) |  |
| Preterm <32 weeks | 3 (15) |
| Preterm 32 to <37 weeks | 4 (20) |
| Term > 37 (37 to <41 weeks) | 13 (65) |
| Birth weight, mean (range) [gram] | 2769 (890 - 4145) |
| Age at time of study, mean (range) [days] | 16.2 (2 - 70) |
| Corrected gestational age at time of study, mean (range) [days] | -3.7 (-30-15) |
| Stay of infant on Neonatal Intensive Care, N (%) |  |
| Yes, up to 1 week | 5 (25) |
| Yes, 1 week or longer | 7 (35) |
| No | 8 (40) |
| Exclusive breastfeeding at time of study, N (%) |  |
| Yes | 7 (35) |
| No, also NG | 9 (45) |
| No, also bottle | 4 (20) |
| Use of nipple shield, N (%) |  |
| For current infant | 9 (45) |
| Only for a previous infant | 1 (5) |
| Never | 10 (50) |

### Vitamin B12 delivery

Observations by the lactation and research team did not report any apparent impact of the tablet’s presence in the nipple shield on the process of feeding. Complete tablet disintegrating and delivery of the full vitamin B12 dose was achieved in all study feeds, with no residue visible to the naked eye. Changes in serum B12 levels are provided in Table 2.

**Table 2:** Changes in serum vitamin B12 levels from baseline to 6-8 hours after the study feed. Data provided are gestational age at birth, and age at time of study. Vitamin B12 levels of four infants were excluded as samples clearly showed haemolysis, affecting the assay’s accuracy.

| Infant ID | Gestational age at birth<br>[weeks+days] | Age at time of study<br>[days from birth] | Baseline serum vitamin B12 level (pre study feed)<br>[pg/mL] | Peak serum vitamin B12 level (6-8 hours post study feed)<br>[pg/mL] | Percentage change |
| --- | --- | --- | --- | --- | --- |
| 1 | 31+5 | 55 | 575 | 3484 | 506 |
| 2 | 26+5 | 70 | 681 | 2577 | 278 |
| 3 | 37+0 | 6 | 449 | 1743 | 288 |
| 4 | 37+2 | 6 | 858 | 1285 | 50 |
| 5 | 41+1 | 4 | 303 | 1045 | 245 |
| 6 | 31+3 | 30 | 236 | 4981 | 2011 |
| 7 | 38+4 | 21 | 593 | 1928 | 225 |
| 8 | 34+6 | 19 | 596 | 4104 | 589 |
| 9 | 41+1 | 6 | 430 | 1006 | 134 |
| 10 | 40+6 | 6 | 925 | 1121 | 21 |
| 11 | 37+6 | 4 | 565 | 1321 | 134 |
| 12 | 41+2 | 6 | 325 | 610 | 88 |
| 13 | 40+2 | 4 | 660 | 1259 | 91 |
| 14 | 40+2 | 6 | 582 | 1104 | 90 |
| 15 | 41+0 | 8 | 397 | 866 | 118 |
| 16 | 32+1 | 29 | 352 | 1506 | 328 |

### Mixed methods assessment

The average duration of semi-structured interviews to assess maternal expectations and experiences was 7.7 min (range 4.4 – 16.6 min) and 7.0 min (range 3.5 - 12.2 min), respectively.

#### Maternal expectations of the study feed

Identified themes related to the uncertainty of risks but also curiosity for the intervention, as well as strong expectations of perceived emotional and practical benefits. Areas of concerns included the use of a nipple shield itself, and the therapeutic it contains. In particular, for mothers who have not previously used nipple shields, unknowns about the infant’s reaction and behaviour during the study feed were the predominant source of worry, including the infant’s ability to latch and the alteration of normal breastfeeding behaviour.

> “It might take some [time] getting used to. I don’t know how she is going to do with a nipple shield. And I don’t know whether it is going to affect her, I mean the way that she latches.” (M12, no NS)

In contrast, mother with previous experience using nipple shields, reflected exclusively on the tablet’s disintegration properties, its potential impact on the milk’s taste and implication on breastfeeding practice.

> “[…] will it dissolve, and will she… be able to taste it?” (M2, NS)
>
> “I don’t have any particular worries. Ok, I suppose if I was going to have a worry it would be that it would give them a negative experience of breastfeeding and then would put them of breastfeeding.” (M6, NS)

Despite associated worries, mothers described their positivity and curiosity in attempting vitamin delivery during breastfeeding.

> “I think it’s worth looking into. It is something I have never thought about. I think it is a good idea.” (M12, no NS)
>
> “It is just quite exciting to see how it works.” (M2, NS)

All participants expected an emotional improvement and increased convenience of infant therapeutic delivery whilst feeding (Table 3). Advantages on an emotional level included the reduction of stress for both mother and infant, as well as a perceived enhancement in physical intimacy for some mothers (Table 3). Perceived practical benefits were related to time saving, and fewer dosing errors using solid dosage forms and being “less messy” (M2, NS). Mothers associated these benefits with breastfeeding as a more ‘natural’ method of delivery compared to oral syringes.

**Table 3:** Comparison of maternal expectations before and experiences reported after the study feed. The Likert scale was used (10 = highly agree, 0 = highly disagree). Further quotes can be found in Table A.1 (supplementary data).

|  | Likert scale<br>before study feed | Likert scale after<br>study feed | Change [%] |
| --- | --- | --- | --- |
| The nipple shield with a vitamin tablet... |  |  |  |
| ...will be / was easier than using an oral syringe. | 7.0 ± 1.6 | 8.3 ± 1.8 | +19 |
| ...will make / made me less worried. | 7.2 ± 2.0 | 8.6 ± 1.5 | +19 |
| ...will make / made my baby feel less upset/ distressed. | 7.7 ± 1.5 | 8.6 ± 1.4 | +12 |
| ...will help / helped me to feel closer to my baby. | 7.7 ± 1.6 | 8.4 ± 1.7 | +9 |

> “It seems a more natural way of administering medication. Because he would have to nurse in this manner. So this shouldn’t be too different from that natural process.” (M5, no NS)

#### Maternal experiences of the study feed

The majority of mothers expressed a positive experience and a feeling of surprise in the nipple shield’s ease of use for therapeutic delivery, and the lack of impact on the infant’s breastfeeding behaviour. This consolidated previous positive thoughts around the use of nipple shields for therapeutic delivery (Table 4). Mothers attributed their surprise about their infants’ contentment to the lack of experience and emotional factors relating to preconceptions about nipple shields.

**Table 4:** Summary of reported maternal experience and acceptability of the study feed. Further quotes can be found in Table A.2 and in Table A.3 (supplementary data).

|  | Agreement [%] |
| --- | --- |
| **Experience** |  |
| The nipple shield with a vitamin tablet... |  |
| ...was a positive experience | 95 |
| ...was easy to use. | 95 |
| ...was comfortable to wear. | 95 |
| My baby.... |  |
| ...latched as usual. | 95 |
| ...breastfed as usual. | 90 |
| **Acceptability** |  |
| I prefer to give medicines / nutrients using a nipple shield over using an oral syringe. | 85 |
| I think the nipple shield could be an acceptable method for <u>nutrient</u> delivery. | 100 |
| I think the nipple shield could be an acceptable method for <u>medicine</u> delivery. | 95 |
| I would like that medicine / nutrient delivery during breastfeeding becomes possible for parents in the future | 100 |

> “I think those worries were probably fear of the unknown. Not ever using a nipple shield before. Sort of remembering how they were three years ago, when I saw them in the shops, and they were a little bit alien-looking… I suppose not having that practice or that experience made me think ‘Oh, what is this going to feel like? And is it going to be a barrier to feeding? And is he going to latch properly.’ But actually, all of that was fine.” (M4, no NS)

Practical concerns remained regarding the potential of incomplete therapeutic delivery.

> “What would you do - if it was an actual drug – and [you had] given only part of a dose? (M2, NS)

#### Overarching themes: perceived advantages and acceptability

Mothers emphasised the potential of this therapeutic delivery method to de-medicalize infant treatment by combining it with the natural process of breastfeeding. It was described by mothers as “less invasive” (M7, no NS), and “not an aggressive method of delivery” (M16, no NS), “you are not forcing them” (M20, no NS). This was seen as a way to ease the emotional burden for mothers of infants with prior neonatal intensive care experience.

> “[…] what’s really nice, especially for [my daughter] - because she started off her life being poked and prodded, and having things stuck in her - that this is such a lovely… like for babies who’ve had to undergo all that, to have something so natural, is lovely.” *(moved to tears)* (M12, no NS)
>
> “[…] for my daughter, because she has been in a hospital for three weeks, I would like something more natural for her from now on. She had - she has - a tube in her nose, and I hope that in the future, we don’t have anything clinical, you know, to deal with. […] the thought [of it] brings us back here. And not that it has been a horrible experience [on the NICU], but it has been very scary. […] at the moment, for us everything that has to do with syringes and medication makes me think of NICU and, you know, this very scary part of her life.” (M19, NS)

All mothers advocated for the availability of oral infant therapeutic administration during breastfeeding, with the majority preferring this method of infant therapeutic delivery (see Table 4), emphasizing that it would provide “choices to make things simpler” (M8, no NS).

> “[…] as parents we have to make sure to give our children medications when they need them, and in the most calm, you know, not upsetting way possible for them. So, having the choice, since every child is different…there isn’t just one way to make it easier, I think. So having more ways means that there will be more children having the best way.” (M19, NS)

## DISCUSSION

This is the first ever proof-of-concept study to demonstrate clinical feasibility and acceptability of therapeutic delivery from a solid formulation during breastfeeding. The study demonstrated that a solid formulation, placed within an ultrathin contact nipple shield, was easily dispersed in human milk and that complete delivery of the tablet’s ‘dose’ was achieved. All infants showed a clear and significant rise in B12 serum levels after their study feed.

The percentage change following B12 delivery varied between infants, and seemed to be neither related to the infants’ gestational age nor to their B12 baseline levels, but may be related to maturational age. Further analyses were beyond the scope of this study, and would require a larger sample size. Future studies on specific formulations would need to assess any inter-individual differences and clinical implications for dosing. The vitamin tablet and nipple shield did not interfere with either infant latch, or maternal breastfeeding experience.

### Maternal perspectives

Although oral syringes are routinely used for the delivery of liquid formulations, maternal responses indicated their association with a predominantly negative, ‘medical’ sensation. While it might be assumed that mothers with previous experience of neonatal intensive care would regard oral syringes as a rather non-invasive intervention, our study showed that nonetheless these mothers perceived the use of oral syringes as stressful.

Despite some pre-conceptions about the use of nipple shields, none of the mothers reported any difficulties with either their use or inclusion of the tablet within the shield. This positive evaluation by mothers on the use of nipple shields contrasts with literature-reported discomfort, inconvenience, and the fear of nipple confusion,[12] but complements positive observations of their benefits in establishing and maintaining breastfeeding with appropriate support.[13,14] Anticipated anxiety related to incomplete delivery, e.g. in case taste could have impacted the infant’s desire to suckle, were not apparent during the study. This aligns with previous literature on the infant’s acquaintance with a variety of tastes and that milk may have taste masking properties.[15–18]

In this setting of a neonatal (intensive) care unit, there was a strong desire to move to a less medicalised approach and gain more ownership in care. This is in keeping with the literature, which suggests mothers welcome an opportunity to build confidence in their own ability to take responsibility for their infant’s care,[19–22] and is an important part of mother-infant-bonding.[23]

### Limitations

This is a single centre study with a limited sample size, intentionally chosen to enable proof-of-concept assessment while reducing unnecessary infant exposure. Studies have not previously investigated single-dose B12 supplementation, and only limited data on pharmacokinetics of B12 in the newborn are available for comparison. Since only mothers willing to use a nipple shield provided their consent, the sample might be biased towards a more favourable evaluation. However, approximately half the participants had not used a nipple shield previously, and only 5% of mothers declined participation due to concerns about its use. Additional studies will be needed to explore different formulations and different age groups.

## CONCLUSIONS

This is the first study to demonstrate the clinical feasibility and acceptability of therapeutic delivery from a solid formulation during breastfeeding. All mothers expressed their support for this approach, with 85% expressing a preference over the use of oral syringes. Further research is warranted to investigate the potential range of therapeutic indications.

## Data Availability

The authors confirm that the data supporting the findings of this study are available within the article and its supplementary material.

## Acknowledgements

The authors like to thank the participants for providing their time to the study. For their support in facilitating the investigation, thanks is given to the nursing and medical teams on both the Neonatal Intensive Care Unit and the Transitional Care Unit at the University of Cambridge Addenbrooke’s Hospital Trust. We would like to thank the Core Biochemical Assay Laboratory (CBAL), Cambridge University Hospitals, for their support in blood sample analysis, and the National Institute for Health Research (NIHR) Cambridge Biomedical Research Centre.

## DECLARATIONS

### Ethics approval

The study was approved by the London, Brighton & Sussex Research Ethics Committee (18/LO/0551). All participants provided their written informed consent to take part and to be quoted anonymously in this publication.

### Competing interests

None declared.

### Funding

The research was supported by a University of Cambridge WD Armstrong PhD studentship, the University of Cambridge Kurt Hahn Trust, and the German Academic Scholarship Foundation.

### Contributors

TM, PP, KB designed the feasibility study and recruitment material. TM led participant recruitment, supported by PP, LB, and KB. TM conducted the interviews and study feeds, supported by PP and LB for several study feeds. TM and KB performed data analysis. SLW provided advice with regard to the study’s mixed methods approach. TM and KB wrote the manuscript, which was revised by PP, LB, SLW, and NKHS. All authors provided approval for publication of the final manuscript.

## Notes

### Competing Interest Statement

The authors have declared no competing interest.

### Clinical Trial

NCT03799367

